# Prevention of liver fibrosis and steatosis progression among heavy drinkers with and without HIV after 30-day drinking-reduction program

**DOI:** 10.1101/2023.02.27.23286532

**Authors:** Seungjun Ahn, Veronica Richards, Emmanuel Thomas, Dushyantha Jayaweera, Varan Govind, Zhigang Li, Ronald A. Cohen, Robert L. Cook

## Abstract

**Background:** This is the first attempt to assess changes in liver abnormalities before and after contingency management (CM) to reduce heavy drinking beyond 30-days of follow-up.

**Objective:** The main objective was to determine whether liver fibrosis and steatosis, measured using FibroScan, change significantly between baseline, 30-days, and 90-days among older adults who drink heavily at baseline, enrolled in the CM intervention for alcohol reduction. The secondary aim of the study was to assess whether the changes in liver outcome measures differ across alcohol consumption categories.

**Methods:** A prospective study (ClinicalTrials.gov registry: NCT03353701) of 46 older adults (63% male, 76.1% Black, mean age = 56.4) with heavy drinking at the baseline, living with or without HIV infection was evaluated. A linear mixed-effects model was used to analyze the FibroScan Transient Elastography (TE for fibrosis) and Controlled Attenuation Parameter (CAP for steatosis).

**Results:** There were no significant changes in liver fibrosis and steatosis measures after 90-days of drinking abstinence among heavy drinkers with or without stratified TE or CAP values at baseline.

**Conclusions:** CM for drinking reduction may not be effective at least short-term prospective in preventing liver fibrosis and steatosis progression for subjects without severe liver disease at baseline.

## Introduction

Alcohol use is the third leading preventable cause of death in the United States [1, 2]. Heavier alcohol use has consistently been reported to be associated with increased risk for numerous chronic diseases and conditions, such as liver diseases [3, 4, 5, 6]. With respect to liver diseases, alcohol-attributable mortality from liver cirrhosis, an advanced stage of liver fibrosis, accounted for 47.9 percent of all liver cirrhosis deaths worldwide (493,300 deaths) [3]. Liver steatosis (fatty liver) is detected in 90% of people with heavy alcohol consumption [5, 7]. Although liver steatosis is usually asymptomatic [5], it can progress to liver inflammation and liver cirrhosis [8]. Though the relationship between heavy alcohol consumption and liver diseases are an established public health concern, it is unclear how reducing or cessating from alcohol consumption is associated with prevention of liver outcomes progression, warranting further investigation.

Alcohol use is prevalent among persons living with HIV (PLWH) [9, 10], which is also associated with incidence of liver disease [9]. Liver disease is one of the leading causes of death among PLWH [10]. A recent cohort study [11] noted that the prevalence of liver diseases among PLWH were 10 times higher compared with HIV-negative counterparts. A systematic review [12] reported that the prevalence of non-alcoholic fatty liver disease (NAFLD) and fibrosis were 35% and 22% in PLWH, respectively. Moreover, HIV is associated with steatosis [13] and chronic systemic inflammation, which may place PLWH at higher risk for developing liver disease [14].

A review article [15] highlighted the need of intervention for drinking reduction and for better understanding of HIV-related health outcomes at individual and policy levels. Contingency management (CM) is one drinking intervention that has been effective for alcohol reduction [16, 17]. CM involves providing financial or other incentives for meeting a treatment goal or, a negative consequence if the individual is unable to meet this goal. However, there is limited published information about how short-term alcohol interventions, such as CM, may impact liver disease, especially in PLWH and in the persons without severe liver disease at baseline. Among two populations of “ healthy” individuals who drank moderately-to-heavily, short-term abstinence (one month) from alcohol resulted in significantly reduced serum alanine aminotransferase (ALT) and gamma-glutamyl transferase (GGT), both indicating improvement in liver function [18, 19]. The study outcomes for these two studies were assessed only at 30-days, so it is unclear if these benefits persisted after 30-days. Further, it is unclear whether any benefits of alcohol reduction extended to prevent liver fibrosis and steatosis progression, key manifestations of both alcoholic-related liver disease (ALD) and NAFLD.

The primary objective of this study was to determine whether liver fibrosis and steatosis, measured using FibroScan, change significantly with alcohol reduction from a CM intervention, over three time periods (baseline, 30-days, and 90-days) among older adults who drink heavily with or without stratified liver fibrosis and steatosis values at baseline. In the current study, the fibrosis-4 (FIB-4) index [20] was analyzed as an additional liver outcome measure. We hypothesized that the patterns of change in liver outcome measures would differ in alcohol consumption categories (*i*.*e. quit/reduced drinking vs. continued heavy drinking*) during the course of study, observed by an interaction effect between alcohol intake and time from an analysis. This study would increase knowledge and empirical evidence on the prevention of progression in liver abnormalities, fibrosis and steatosis before and after the initiation of a drinking abstinence program.

## Materials and Methods

### Study Participants

This prospective study included older adults (aged 50 to 75 years) with heavy drinking (≥14 drinks/week for women, ≥21 drinks/week for men) at the baseline, living with or without HIV infection, and were enrolled in a clinical trial of drinking-reduction from 12/11/2017 to 11/16/2021. A description of the methods of drinking-reduction program, “ 30-Day Challenge Study,” is available in ClinicalTrials.gov registry and is shown in Fig 1. The trial was registered under ClinicalTrials.gov with registry date and number (11/27/2017) (NCT03353701) respectively. In brief, the “ 30-Day Challenge Study” used CM with financial incentives to measure the impact of changes in alcohol consumption on changes in neurocognitive disorders, liver fibrosis, and other HIV-related comorbidities. All procedures in the “ 30-Day Challenge Study” were approved by the central Institutional Review Board at University of Miami (Study Number: 20170396). All study participants provided informed consent prior to participation. All methods were carried out in accordance with relevant guidelines and regulations.

**Fig 1.**
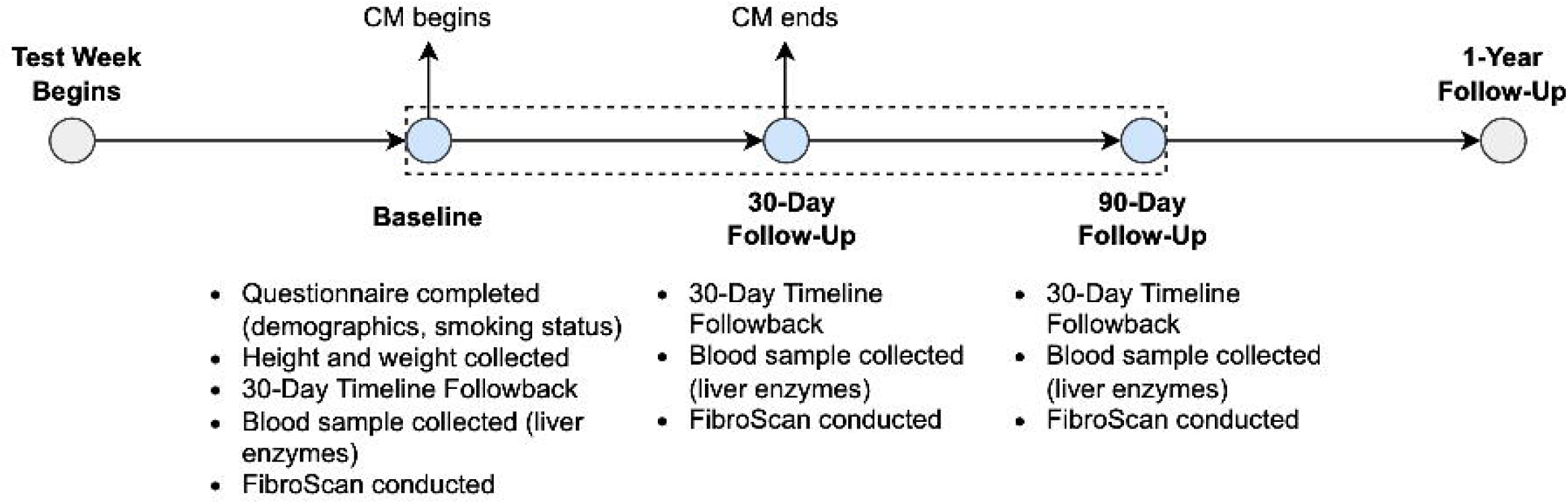
Timeline of the 30-Day Challenge Study. The data used in this analysis come from the baseline, 30-day, and 90-day follow-up visits, with measures included listed below each timepoint. The contingency management (CM) period began at Baseline and ended at the 30-day follow-up.

This study analyzed data from the baseline, 30-day follow-up (post CM), and 90-day follow-up visits. At the time of data retrieval (12/07/2021), the dataset included 56 participants, but 10 were excluded from the analysis due to missing baseline (*n* = 1), or missing both 30-days and 90-days follow-up data on alcohol consumption (*n* = 8), or AST level (*n* = 1). In summary, data were included from 46 participants with complete information on alcohol intake and liver outcome measures at baseline and at least one follow-up visit.

### Outcome Measures

The outcome variables included the changes from baseline values on liver fibrosis measured by transient elastography (TE), and steatosis measured by controlled attenuation parameter (CAP), respectively. These quantitative measures were obtained using a FibroScan (Echosens, Paris, France) from baseline, 30-days, and 90-days. Additionally, the fibrosis-4 (FIB-4) index [20], a validated noninvasive test alternative to liver biopsy, was considered to further investigate the trend in liver fibrosis, calculated as,

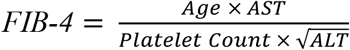

where AST = aspartate aminotransferase and ALT = alanine aminotransferase. Hence, the changes from baseline value on FIB-4 was also considered as an outcome variable.

### Main Predictor

Although abstinence is often regarded as the primary endpoint in alcohol use disorder treatment, the prior studies [21, 22] have shown that reduced alcohol consumption at any level has also been linked to improved health outcomes. Thus, we decided to examine alcohol consumption as a dichotomous variable rather than a 3-level categorical variable. Alcohol intake categories were derived from the Alcohol Timeline Followback (TLFB) [23] data in the previous 30-days. *Heavy drinking* was defined as consuming more than 7 drinks per week for females and more than 14 drinks per week for males, based on the National Institute on Alcohol Abuse and Alcoholism (NIAAA) guideline [24]; *reduced drinking* was definined as drinking level that did not qualify as heavy drinking; 3) *quit drinking* if a participant did not drink during 30 days. In this study, reduced and quit drinking categories are combined for the analysis.

### Covariates

HIV diagnosis (negative or positive), body mass index (BMI), sex at birth (female or male), age in continuous scale, and hepatitis C (HCV) were included as covariates in later multivariable analysis. HCV antibody test was conducted at baseline and classified as positive or negative. HCV was included as a covariate, as earlier research showed that the alcohol consumption is related to the transmission of both HIV and HCV [25]. BMI was broken down into three categories: underweight or normal (BMI < 25.0), overweight (25.0 ≤ BMI < 30.0), and obese (BMI ≥ 30) based on the Centers for Disease Control and Prevention classification of overweight and obesity guideline [26]. Previous studies have shown that obesity is the important risk factor for NAFLD [8, 27].

### Statistical Methods

A linear mixed-effects model with random intercept was used to determine the changes in FibroScan (TE and CAP), and FIB-4 score while accounting for within-subject correlations. An unstructured covariance matrix was used. The change in log-transformed values of TE and FIB-4 were used as outcomes after examining residual diagnostics and standard model assumptions (normality). However, the results were back-transformed and presented in their original scales for the purpose of simplicity. As a subgroup analysis, we performed similar analyses among participants with abnormal baseline values of TE (≥ 7 kPa) and CAP (≥ 238 dB/m) [28, 29].

We firstly fitted the marginal models (unadjusted models) for each predictor variable to identify covariates that are marginally associated with each liver outcome variable at *p* < 0.10 (forward selection). Subsequently, multivariable models (adjusted models) included time, categorical alcohol intake, interaction term with alcohol intake and time, and the covariates that were significantly associated with each liver outcome variables from unadjusted models. Lastly, backward elimination was applied to identify covariates that are associated with liver outcome variables in adjusted models, including only covariates with *p* < 0.05. Time was treated as a continuous variable.

Demographic characteristics were presented as mean and standard deviation (SD) for continuous variables and frequencies with percentages for categorical variables. Results were considered statistically significant when *p* < 0.05. All analyses were conducted using SAS version 9.4 (SAS Institute, Inc., Cary, NC).

### Sample Size Considerations

The sample size (*n* = 46) was based on availability of data during the time span of the database retrieval. A sample size of 46 yielded 85.7% statistical power to detect a large effect size (Cohen’s *f* = 0.5), determined by repeated measures ANOVA with a significance level of 0.05. A statistical power calculation was performed in R version 4.0.2 (R Foundation for Statistical Computing, Vienna, Austria).

## Results

### Demographic and Clinical Characteristics

The baseline characteristics are summarized in Table 1. The final data included 46 participants. Overall, the mean age of study participants was 56.4 years (SD = 4.7 years). A majority of participants were diagnosed with HIV (*n =* 28; 60.9%) and were either overweight (*n =* 15; 32.6%) or obese (*n =* 13; 28.3%). Most participants were Black (*n =* 35; 76.1%) and male sex (*n =* 29; 63.0%). With 9 and 3 missing values, a majority of participants smoked cigarettes (*n =* 30; 81.1%) and were HCV negative (*n =* 38; 88.4%) at the time of enrollment in the 30-Day Challenge Study.

**Table 1.**
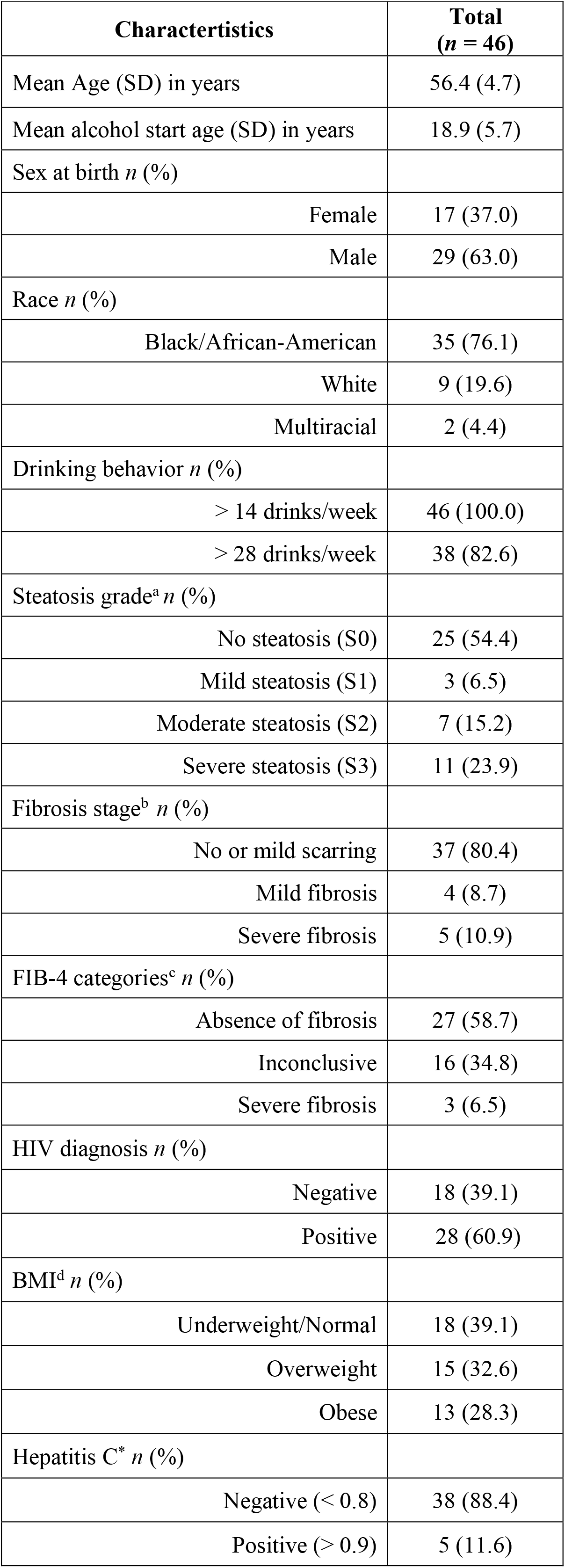
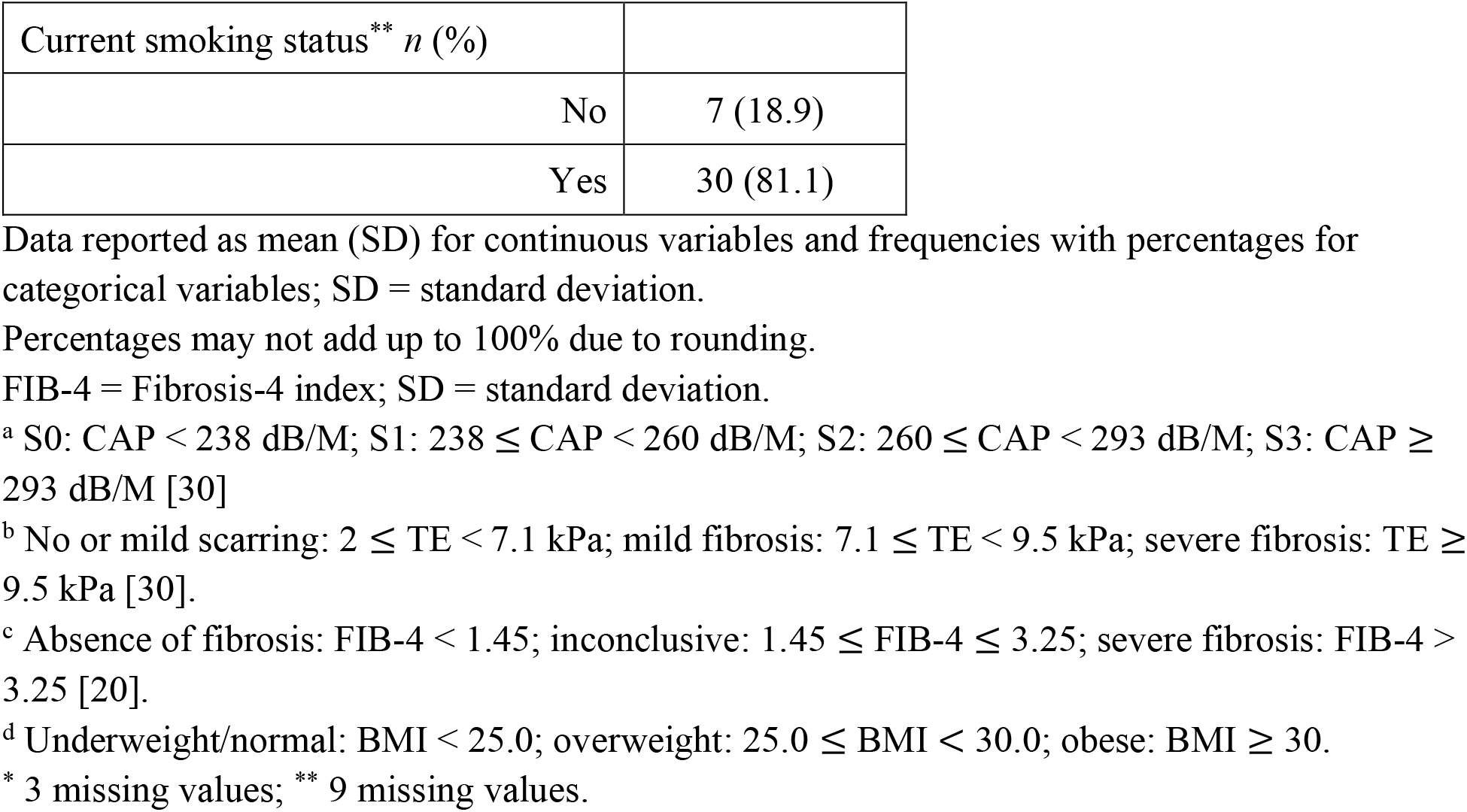
Baseline characteristics of study participants in the study.

Approximately 20% of participants had mild (*n* = 4) or severe (*n* = 5) fibrosis and almost half (n = 21; 45.6%) had steatosis. The majority of participants did not have fibrosis (*n* = 27; 58.7%) based on FIB-4 index.

### Unadjusted Models for Screening Covariates (Forward Selection of Model Covariates)

In the unadjusted model using the entire sample, age (type III *p* = 0.047) and sex (type III *p* = 0.019) were marginally associated with the the change in CAP. None of the covariates were significantly associated with changes in TE or FIB-4.

In the marginal analysis using subsamples with abnormal baseline TE, HCV (type III *p =* 0.010) and sex (type III *p =* 0.015) were significantly associated with change in TE. There were marginal associations between FIB-4 and HCV (type III *p =* 0.080). None of covariates were significantly associated with change in CAP in this subsample.

With subsamples of abnormal baseline CAP, age (type III *p =* 0.009), HIV (type III *p =* 0.052), and BMI (type III *p =* 0.012) were significantly associated with change in CAP. There was significant marginal association between FIB-4 and BMI (type III *p* = 0.009). Lastly, TE did not have marginal association with any covariates.

Subsequently, marginally significant covariates were included in the initial multivariable model for the backward elimination.

### Adjusted Models for Liver Outcomes Using Entire Samples (*n* = 46)

The results presented in Table 2 indicate that there was no significant change in any of liver outcomes for every one-day increase in time regardless of the level of alcohol intake (interaction terms). Males had significantly smaller changes in levels of CAP (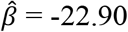; 95% CI of 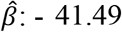, -4.301; *p* = 0.017), when compared to females. Older age was associated with greater changes in levels of CAP (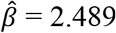; 95% CI of 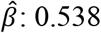, 4.441; *p* = 0.014).

**Table 2.** Changes in liver outcomes associated with changes in alcohol consumption at 30 and 90 days among 46 adults participating in a contingency management study.

| Variables | Outcomes |  |  |  |  |  |  |  |  |
| --- | --- | --- | --- | --- | --- | --- | --- | --- | --- |
|  | Change in TE |  |  | Change in CAP |  |  | Change in FIB-4 |  |  |
| | exp ($\hat{\beta}$) | 95% CI of exp ($\hat{\beta}$) | p-value | $\hat{\beta}$ | 95% CI of $\hat{\beta}$ | p-value | exp ($\hat{\beta}$) | 95% CI of exp ($\hat{\beta}$) | p-value |
| **Main Predictors** |  |  |  |  |  |  |  |  |  |
| Time | 1.000 | 0.997, 1.002 | 0.930 | 0.022 | -0.321, 0.366 | 0.895 | 1.000 | 0.997, 1.003 | 0.913 |
| Alcohol intake |  |  |  |  |  |  |  |  |  |
| Continued heavy drinking | ‡ | ‡ | ‡ | ‡ | ‡ | ‡ | ‡ | ‡ | ‡ |
| Quit/Reduced drinking | 1.080 | 0.849, 1.372 | 0.521 | -22.96 | -63.31, 17.40 | 0.255 | 1.053 | 0.795, 1.393 | 0.712 |
| Time × I(Alcohol intake = Quit/Reduced drinking) | 0.999 | 0.995, 1.002 | 0.485 | 0.343 | -0.244, 0.931 | 0.243 | 0.999 | 0.995, 1.004 | 0.810 |
| Baseline liver measure<sup>a</sup> | 0.786 | 0.633, 0.977 | 0.031* | -0.246 | -0.420, -0.072 | 0.007* | 0.674 | 0.502, 0.905 | 0.010* |
| **Covariates<sup>b</sup>** |  |  |  |  |  |  |  |  |  |
| Age |  |  |  | 2.489 | 0.538, 4.441 | 0.014* |  |  |  |
| Sex at birth |  |  |  |  |  |  |  |  |  |
| Female |  |  |  | ‡ | ‡ | ‡ |  |  |  |
| Male |  |  |  | -22.90 | -41.49, -4.301 | 0.017* |  |  |  |

All mixed-effects models included linear function of time, categorical alcohol intake, and an interaction term with categorical alcohol intake and time. An unstructured covariance structure was used.
The coefficient estimates for TE and FIB-4 were back-transformed, that is, exponent of the coefficient estimates.
<sup>a</sup> This is to adjust the outcome at baseline. For example, CAP at baseline is used as a predictor for the fitted model for change in CAP as an outcome variable.
<sup>b</sup> In these models, age in years, HIV diagnosis, BMI categories, hepatitis C diagnosis, and sex were firstly considered as potential confounders and to be selected from unadjusted models at the 0.1 significance level. Selected covariates were then remained after backward elimination at the 0.05 significance level.
TE = fibrosis measured using FibroScan; CAP = steatosis measured using FibroScan; FIB-4 = Fibrosis-4 index; $\hat{\beta}$ = coefficient estimates; CI = confidence interval
\* A p-value < 0.05 was considered statistically significant, unless otherwise noted.
‡ Reference group for comparisons.

### Adjusted Models for Liver Outcomes with Abnormal Baseline TE Samples (*n* = 9)

The interaction terms of the mixed-effects model results using samples with abnormal baseline TE (Table 3) highlights that there were no significant changes in liver outcome measures with varying alcohol intake categories for every one-day increase in time. Of them, the greater change (increase) in FIB-4 from baseline for every one-day increase in time was observed for those who quit or reduced drinking 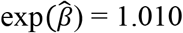 95% CI of 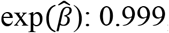): 0.999, 1.021; *p* = 0.067), when compared with continued heavy drinking, albeit not statistically significant. This implies that 34.8% higher level of FIB-4 in quit or reduced drinking versus continued heavy drinking after 30 days, calculated by 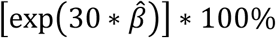.

**Table 3.**
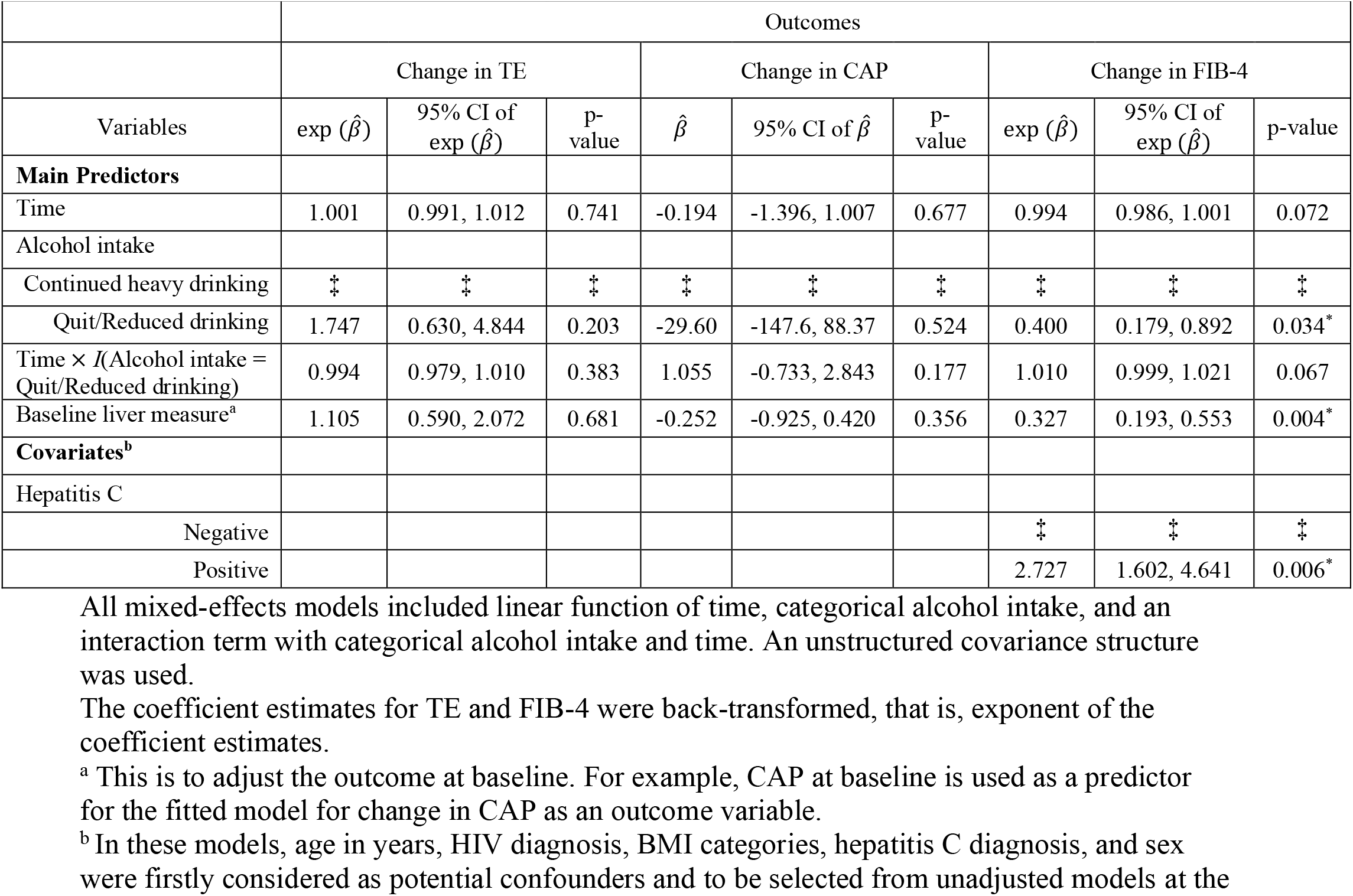
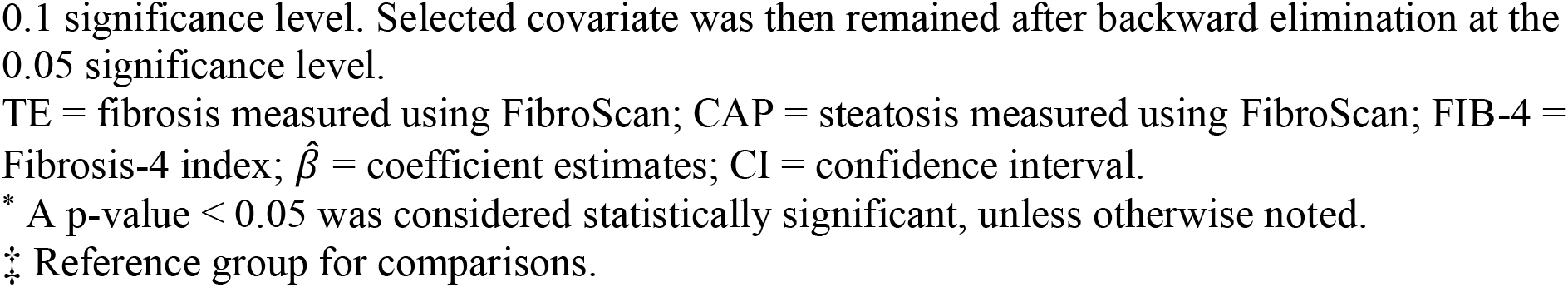
Changes in liver outcomes associated with changes in alcohol consumption at 30 and 90 days among 9 adults with abnormal baseline TE (TE ≥ 7) participating in a contingency management study.

Sex (*p* = 0.229) and HCV (*p* = 0.086) were excluded from the final adjusted model after the backward elimination for TE. HCV was remained significant in the model for FIB-4 (*p* = 0.006) after the backward elimination.

### Adjusted Models for Liver Outcomes with Abnormal Baseline CAP Samples (*n* = 21)

In the mixed-effects model using samples with abnormal baseline CAP (Table 4), there was no statistically significant interaction effect of time and alcohol intake categories on changes in liver outcome measures. Although it was not statistically significant, it is noteworthy that the greater change (increase) in CAP from baseline for every one-day increase in time was observed for those who quit or reduced drinking (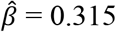; 95% CI of 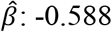, 1.218; *p* = 0.465), when compared with continued heavy drinking.

**Table 4.**
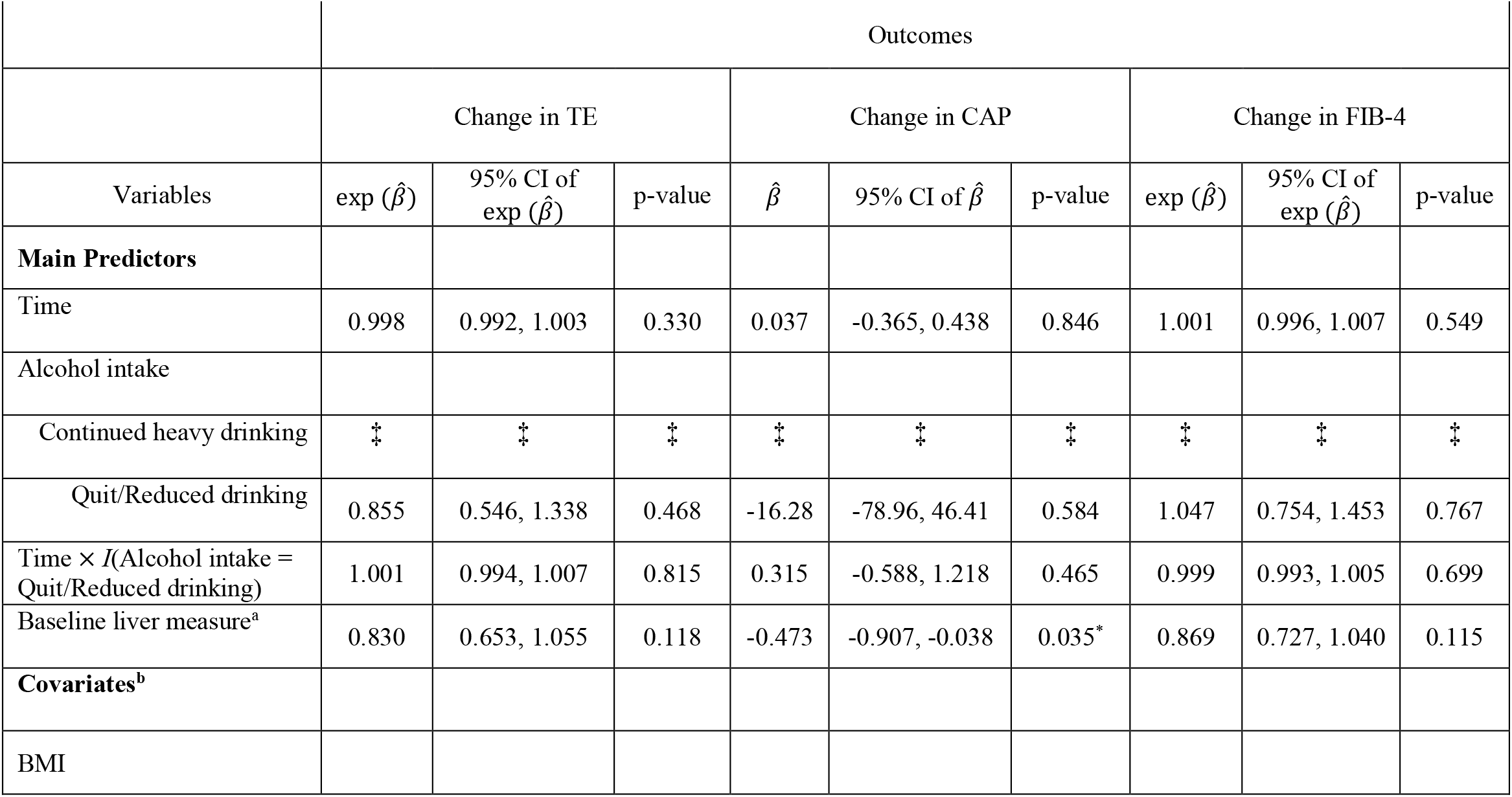
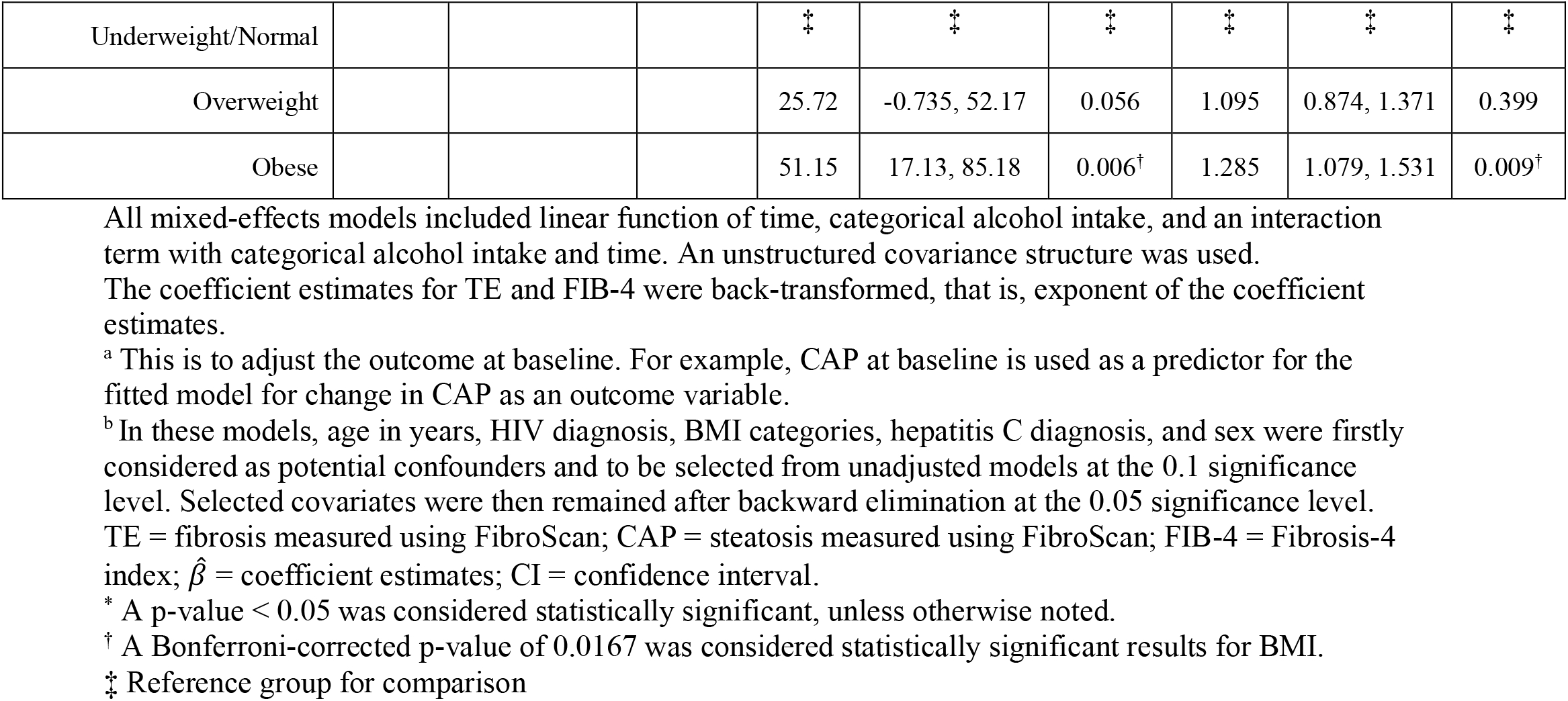
Changes in liver outcomes associated with changes in alcohol consumption at 30 and 90 days among 21 adults with abnormal baseline CAP (CAP ≥ 238) participating in a contingency management study.

|  | Outcomes |  |  |  |  |  |  |  |  |
| --- | --- | --- | --- | --- | --- | --- | --- | --- | --- |
|  | Change in TE |  |  | Change in CAP |  |  | Change in FIB-4 |  |  |
| Variables | exp ($\hat{\beta}$) | 95% CI of exp ($\hat{\beta}$) | p-value | $\hat{\beta}$ | 95% CI of $\hat{\beta}$ | p-value | exp ($\hat{\beta}$) | 95% CI of exp ($\hat{\beta}$) | p-value |
| Main Predictors |  |  |  |  |  |  |  |  |  |
| Time | 0.998 | 0.992, 1.003 | 0.330 | 0.037 | -0.365, 0.438 | 0.846 | 1.001 | 0.996, 1.007 | 0.549 |
| Alcohol intake |  |  |  |  |  |  |  |  |  |
| Continued heavy drinking | ‡ | ‡ | ‡ | ‡ | ‡ | ‡ | ‡ | ‡ | ‡ |
| Quit/Reduced drinking | 0.855 | 0.546, 1.338 | 0.468 | -16.28 | -78.96, 46.41 | 0.584 | 1.047 | 0.754, 1.453 | 0.767 |
| Time × I(Alcohol intake = Quit/Reduced drinking) | 1.001 | 0.994, 1.007 | 0.815 | 0.315 | -0.588, 1.218 | 0.465 | 0.999 | 0.993, 1.005 | 0.699 |
| Baseline liver measure<sup>a</sup> | 0.830 | 0.653, 1.055 | 0.118 | -0.473 | -0.907, -0.038 | 0.035* | 0.869 | 0.727, 1.040 | 0.115 |
| Covariates<sup>b</sup> |  |  |  |  |  |  |  |  |  |
| BMI |  |  |  |  |  |  |  |  |  |
| Underweight/Normal |  |  |  | ‡ | ‡ | ‡ | ‡ | ‡ | ‡ |
| Overweight |  |  |  | 25.72 | -0.735, 52.17 | 0.056 | 1.095 | 0.874, 1.371 | 0.399 |
| Obese |  |  |  | 51.15 | 17.13, 85.18 | 0.006† | 1.285 | 1.079, 1.531 | 0.009† |

All mixed-effects models included linear function of time, categorical alcohol intake, and an interaction term with categorical alcohol intake and time. An unstructured covariance structure was used. The coefficient estimates for TE and FIB-4 were back-transformed, that is, exponent of the coefficient estimates. <sup>a</sup> This is to adjust the outcome at baseline. For example, CAP at baseline is used as a predictor for the fitted model for change in CAP as an outcome variable. <sup>b</sup> In these models, age in years, HIV diagnosis, BMI categories, hepatitis C diagnosis, and sex were firstly considered as potential confounders and to be selected from unadjusted models at the 0.1 significance level. Selected covariates were then remained after backward elimination at the 0.05 significance level. TE = fibrosis measured using FibroScan; CAP = steatosis measured using FibroScan; FIB-4 = Fibrosis-4 index; $\hat{\beta}$ = coefficient estimates; CI = confidence interval. \* A p-value < 0.05 was considered statistically significant, unless otherwise noted. † A Bonferroni-corrected p-value of 0.0167 was considered statistically significant results for BMI. ‡ Reference group for comparison

BMI remained in the final adjusted model for CAP and FIB-4 after backward elimination. Obese (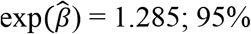 CI of 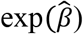: 1.079, 1.531; *p* = 0.009) participants had 28.5% significantly higher level of FIB-4 from baseline, comparing with those who were either underweight or normal. Lastly, HIV (*p* = 0.174) and age (*p =* 0.066) were excluded from the final adjusted models for CAP after the backward elimination.

## Discussion

To our knowledge, this study is the first of its kind in the literature to evaluate liver abnormalities before and after CM to reduce heavy drinking beyond 30-days of follow-up. To date, no studies have examined whether reduction in drinking will have an effect in a population of persons without severe liver disease to start with, but who are at high risk for progression. In this study, the interaction term obtained in an analysis showed that there were no significant changes in liver abnormalities after 90-days of drinking abstinence or reduction among heavy drinkers with or without stratified TE and CAP values at baseline. This suggests that CM for drinking reduction may not be effective at least short-term prospective in improving liver fibrosis and steatosis for subjects with or without abnormal levels of TE or CAP. It is not surprising to see that the behavioral intervention has not always been successful in improving health outcomes. A recent study has shown that the dietary behavioral intervention to increase vegetable intake did not significantly reduce cancer progression in men with early-stage prostate cancer [31]. In addition, results of the meta-analysis suggested that a brief intervention on alcohol consumption did not substantially reduce cigarette smoking in terms of frequency of smoking and likelihood of smoking cessation [32]. Although we did not see an improvement in our study, it is noteworthy that there was no significant change in liver abnormalities over time. Therefore, the liver disease did not progress further stages. It remains a question whether our intervention is what prevented the disease progression.

Additional findings of this study have identified a number of covariates that are significantly associated with liver outcome measures. Participants with HCV infection had substantially higher levels of FIB-4 with stratified TE values at baseline, when compared with those without HCV infection. It is important to note that none of the participants had advanced alcoholic liver disease or acute alcoholic hepatitis at the time of study enrollment. Moreover, the majority of the study participants were recruited from an HIV clinic, not a liver clinic. Thus, we may have less opportunities to view improvement in liver outcomes.

This study has a number of limitations to be considered. First, the alcohol intake categories variable was created based on the TLFB method to calculate average number of drinks per week. A recall bias and social desirability bias in self-reported longitudinal number of drinks per day may lead to an inaccurate estimation of parameter estimates of the model. This may be especially true during the CM period, as contingencies were aligned with drinking abstinence. Second, although our results will contribute future understanding on changes in liver outcome measures, the fitted model was constructed using a dataset with a limited number of covariates and relatively small sample size.

Future studies with a larger sample size and diverse cohort characteristics (*e*.*g*. adjusted FibroScan scores by GGT and lifestyle variable such as health satisfaction or well-being) are warranted to validate our findings.

## Data Availability

All data produced in the present study are available upon reasonable request to the authors.

## Acknowledgments

We are grateful for the administrative support of Dr. Christine Frank and for the data management support of Dr. Zhi Zhou and Mr. Alex Li at the Southern HIV and Alcohol Research Consortium.

## Abbreviations

CM: contingency management;
TE: Transient elastography;
CAP: Controlled attenuation parameter;
HIV: Human immunodeficiency virus;
PLWH: Persons living with HIV;
NAFLD: Non-alcoholic fatty liver disease;
ALT: aminotransferase;
GGT: gamma-glutamyl transferase;
ALD: Alcoholic-related liver disease;
FIB-4: Fibrosis-4;
AST: Aspartate aminotransferase;
TLFB: Alcohol timeline followback;
HCV: Hepatitis C

## Supporting Information

**S1 Table. Crude descriptive statistics of liver outcome measures at baseline, 30-days, and 90-days.**

**S2 Table. Descriptive statistics of change in liver outcome measures at 30-days and 90-days with quit/reduced and continued heavy drinking.**

**S3 Table. Changes in liver outcomes associated with changes in alcohol consumption at 30 and 90 days among 46 adults participating in a contingency management study.**

**S4 Table. Changes in liver outcomes associated with changes in alcohol consumption at 30 and 90 days among 9 adults with abnormal baseline TE (TE ≥ 7) participating in a contingency management study.**

**S5 Table. Changes in liver outcomes associated with changes in alcohol consumption at 30 and 90 days among 21 adults with abnormal baseline CAP (CAP ≥ 238) participating in a contingency management study.**

