## Supplementary Tables S1 to S5 for "Prevention of liver fibrosis and steatosis progression among heavy drinkers with and without HIV after 30-day drinking-reduction program"

### Supplementary Information

The unadjusted summary statistics of each liver outcome measured at the baseline, 30-day follow-up (post CM), and 90-day follow-up visits are outlined in S1 Table. S2 Table summarizes the mean change in each liver outcome measures in those who quit/reduced, or continued heavy drinking at 30-days and 90-days. S3-S5 Tables summarize results of final adjusted models including binary time (30 or 90-days), change in weekly alcohol intake in continuous scale, and an interaction term with change in weekly alcohol intake and binary time, respectively.

**S1 Table. Crude descriptive statistics of liver outcome measures at baseline, 30-days, and 90-days.**

| **Liver Outcomes** | **Baseline (*n* = 46)** | **30-Days (*n* = 44)** | **90-Days (*n* = 39)** |
|---|---|---|---|
| TE | 6.0 (4.1) | 6.0 (3.8) | 5.8 (4.1) |
| CAP | 245.7 (50.3) | 234.3 (54.0) | 245.9 (51.6) |
| FIB-4 | 1.6 (1.2) | 1.4 (0.8) | 1.4 (0.9) |

Data reported as mean (SD) for continuous variables; SD = standard deviation.
TE = fibrosis measured using FibroScan; CAP = steatosis measured using FibroScan; FIB-4 = Fibrosis-4 index.

**S2 Table. Descriptive statistics of change in liver outcome measures at 30-days and 90-days with quit/reduced and continued heavy drinking.**

| **Change in Liver Outcomes** | **30-Days (*n* = 44)** | | **90-Days (*n* = 39)** | |
|---|---|---|---|---|
| | Continued heavy drinking (*n* = 24) | Quit/Reduced drinking (*n* = 20) | Continued heavy drinking (*n* = 17) | Quit/Reduced drinking (*n* = 22) |
| TE | -0.3 (2.3) | 0.1 (2.0) | -0.2 (1.6) | -0.3 (1.5) |
| CAP | -2.8 (36.9) | -20.1 (48.6) | -6.2 (41.6) | 2.9 (47.9) |
| FIB-4 | -0.3 (1.3) | -0.2 (0.4) | -0.2 (0.4) | -0.1 (0.7) |

Data reported as mean (SD) for continuous variables; SD = standard deviation.
TE = fibrosis measured using FibroScan; CAP = steatosis measured using FibroScan; FIB-4 = Fibrosis-4 index.

**S3 Table. Changes in liver outcomes associated with changes in alcohol consumption at 30 and 90 days among 46 adults participating in a contingency management study.**

| | Outcomes | | | | | | | | |
|---|---|---|---|---|---|---|---|---|---|
| | Change in TE | | | Change in CAP | | | Change in FIB-4 | | |
| Variables | exp ($\hat{\beta}$) | 95% CI of exp ($\hat{\beta}$) | p-value | $\hat{\beta}$ | 95% CI of $\hat{\beta}$ | p-value | exp ($\hat{\beta}$) | 95% CI of exp ($\hat{\beta}$) | p-value |
| **Main Predictors** | | | | | | | | | |
| Time | | | | | | | | | |
| 30-Days | ‡ | ‡ | ‡ | ‡ | ‡ | ‡ | ‡ | ‡ | ‡ |
| 90-Days | 0.936 | 0.800, 1.095 | 0.398 | -3.866 | -30.67, 22.93 | 0.771 | 0.997 | 0.838, 1.186 | 0.973 |
| Change in weekly alcohol intake | 1.002 | 0.999, 1.005 | 0.141 | 0.233 | -0.215, 0.681 | 0.298 | 1.003 | 1.000, 1.006 | 0.069 |
| *I*(Time = 90-Days) × Alcohol intake | 1.000 | 0.997, 1.002 | 0.910 | -0.330 | -0.796, 0.137 | 0.160 | 1.001 | 0.998, 1.004 | 0.687 |
| Baseline liver measure[1] | 0.754 | 0.589, 0.964 | 0.026 | -0.246 | -0.426, -0.066 | 0.009* | 0.654 | 0.475, 0.899 | 0.011* |
| Baseline weekly alcohol intake | 1.004 | 1.000, 1.008 | 0.083 | -0.132 | -0.570, 0.307 | 0.545 | 1.005 | 1.001, 1.008 | 0.013 |
| **Covariates[2]** | | | | | | | | | |
| Age | | | | 2.394 | 0.291, 4.497 | 0.027* | | | |
| Sex at birth | | | | | | | | | |
| Female | | | | ‡ | ‡ | ‡ | | | |
| Male | | | | -20.68 | -38.79, -2.581 | 0.026* | | | |

All models included binary time (30 or 90-days), change in weekly alcohol intake in continuous scale, and an interaction term with change in weekly alcohol intake and binary time. An unstructured covariance structure was used.

The coefficient estimates for TE and FIB-4 were back-transformed, that is, exponent of the coefficient estimates.

[1] This is to adjust the outcome at baseline. For example, CAP at baseline is used as a predictor for the fitted model for change in CAP as an outcome variable.

[2] In these models, age in years, HIV diagnosis, BMI categories, hepatitis C diagnosis, and sex were considered as potential confounders and to be selected from unadjusted models at the 0.1 significance level. Selected covariates were then remained after backward elimination at the 0.05 significance level.

TE = fibrosis measured using FibroScan; CAP = steatosis measured using FibroScan; FIB-4 = Fibrosis-4 index; $\hat{\beta}$ = coefficient estimates; CI = confidence interval.

* A p-value < 0.05 was considered statistically significant, unless otherwise noted.

‡ Reference group for comparisons

**S4 Table. Changes in liver outcomes associated with changes in alcohol consumption at 30 and 90 days among 9 adults with abnormal baseline TE (TE ≥ 7) participating in a contingency management study.**

| | Outcomes | | | | | | | | |
|---|---|---|---|---|---|---|---|---|---|
| | Change in TE | | | Change in CAP | | | Change in FIB-4 | | |
| Variables | exp ($\hat{\beta}$) | 95% CI of exp ($\hat{\beta}$) | p-value | $\hat{\beta}$ | 95% CI of $\hat{\beta}$ | p-value | exp ($\hat{\beta}$) | 95% CI of exp ($\hat{\beta}$) | p-value |
| **Main Predictors** | | | | | | | | | |
| Time | | | | | | | | | |
| 30-Days | ‡ | ‡ | ‡ | ‡ | ‡ | ‡ | ‡ | ‡ | ‡ |
| 90-Days | 0.737 | 0.294, 1.847 | 0.409 | -85.22 | -203.9, 33.47 | 0.117 | 0.872 | 0.326, 2.328 | 0.718 |
| Change in weekly alcohol intake | 1.005 | 0.991, 1.109 | 0.380 | -0.122 | -1.688, 1.443 | 0.839 | 1.009 | 0.995, 1.023 | 0.152 |
| *I*(Time = 90-Days) × Alcohol intake | 0.997 | 0.983, 1.011 | 0.587 | -1.697 | -3.507, 0.113 | 0.060 | 0.999 | 0.984, 1.014 | 0.796 |
| Baseline liver measure[1] | 1.031 | 0.354, 3.002 | 0.940 | -0.273 | -0.827, 0.280 | 0.242 | 0.385 | 0.209, 0.712 | 0.013* |
| Baseline weekly alcohol intake | 1.008 | 0.984, 1.032 | 0.424 | -0.311 | -2.484, 1.861 | 0.711 | 1.012 | 0.991, 1.032 | 0.186 |
| **Covariates**[2] | | | | | | | | | |
| Hepatitis C | | | | | | | | | |
| Negative | | | | | | | ‡ | ‡ | ‡ |
| Positive | | | | | | | 2.333 | 1.219, 4.462 | 0.022* |

All models included binary time (30 or 90-days), change in weekly alcohol intake in continuous scale, and an interaction term with change in weekly alcohol intake and binary time. An unstructured covariance structure was used.

The coefficient estimates for TE and FIB-4 were back-transformed, that is, exponent of the coefficient estimates.

[1] This is to adjust the outcome at baseline. For example, CAP at baseline is used as a predictor for the fitted model for change in CAP as an outcome variable.

[2] In these models, age in years, HIV diagnosis, BMI categories, hepatitis C diagnosis, and sex were considered as potential confounders and to be selected from unadjusted models at the 0.1 significance level. Selected covariates were then remained after backward elimination at the 0.05 significance level.

TE = fibrosis measured using FibroScan; CAP = steatosis measured using FibroScan; FIB-4 = Fibrosis-4 index; $\hat{\beta}$ = coefficient estimates; CI = confidence interval.

* A p-value < 0.05 was considered statistically significant, unless otherwise noted.

‡ Reference group for comparisons.

**S5 Table. Changes in liver outcomes associated with changes in alcohol consumption at 30 and 90 days among 21 adults with abnormal baseline CAP (CAP ≥ 238) participating in a contingency management study.**

| | Outcomes | | | | | | | | |
|---|---|---|---|---|---|---|---|---|---|
| | Change in TE | | | Change in CAP | | | Change in FIB-4 | | |
| Variables | exp ($\hat{\beta}$) | 95% CI of exp ($\hat{\beta}$) | p-value† | $\hat{\beta}$ | 95% CI of $\hat{\beta}$ | p-value† | exp ($\hat{\beta}$) | 95% CI of exp ($\hat{\beta}$) | p-value† |
| **Main Predictors** | | | | | | | | | |
| Time | | | | | | | | | |
| 30-Days | ‡ | ‡ | ‡ | ‡ | ‡ | ‡ | ‡ | ‡ | ‡ |
| 90-Days | 0.722 | 0.621, 0.840 | 0.0004* | -17.19 | -71.59, 37.22 | 0.507 | 0.937 | 0.809, 1.085 | 0.352 |
| Change in weekly alcohol intake | 1.006 | 1.004, 1.009 | < 0.0001* | 0.427 | -0.353, 1.208 | 0.258 | 1.005 | 1.002, 1.007 | 0.0007* |
| *I*(Time = 90-Days) × Alcohol intake | 0.997 | 0.995, 0.999 | 0.008* | -0.624 | -1.478, 0.229 | 0.138 | 0.999 | 0.996, 1.001 | 0.323 |
| Baseline liver measure[1] | 0.804 | 0.607, 1.064 | 0.117 | -0.563 | -0.974, -0.153 | 0.011* | 0.888 | 0.744, 1.059 | 0.169 |
| Baseline weekly alcohol intake | 1.006 | 0.999, 1.012 | 0.082 | 0.479 | -0.326, 1.284 | 0.221 | 1.005 | 1.000, 1.010 | 0.043* |
| **Covariates[2]** | | | | | | | | | |
| BMI | | | | | | | | | |
| Underweight/Normal | | | | ‡ | ‡ | ‡ | ‡ | ‡ | ‡ |
| Overweight | | | | 22.60 | -1.179, 46.37 | 0.061 | 1.103 | 0.912, 1.333 | 0.285 |
| Obese | | | | 60.34 | 33.82, 86.86 | 0.0003† | 1.327 | 1.051, 1.675 | 0.021 |

All models included binary time (30 or 90-days), change in weekly alcohol intake in continuous scale, and an interaction term with change in weekly alcohol intake and binary time. An unstructured covariance structure was used.

The coefficient estimates for TE and FIB-4 were back-transformed, that is, exponent of the coefficient estimates.

[1] This is to adjust the outcome at baseline. For example, CAP at baseline is used as a predictor for the fitted model for change in CAP as an outcome variable.

[2] In these models, age in years, HIV diagnosis, BMI categories, hepatitis C diagnosis, and sex were considered as potential confounders and to be selected from unadjusted models at the 0.1 significance level. Selected covariates were then remained after backward elimination at the 0.05 significance level.

TE = fibrosis measured using FibroScan; CAP = steatosis measured using FibroScan; FIB-4 = Fibrosis-4 index; $\hat{\beta}$ = coefficient estimates; CI = confidence interval.

* A p-value < 0.05 was considered statistically significant, unless otherwise noted.

† A Bonferroni-corrected p-value of 0.0167 was considered statistically significant results for BMI.

‡ Reference group for comparisons.
